# Prevalence, treatment, and factors associated with cryptococcal meningitis post introduction of integrase inhibitors antiretroviral based regimens among people living with HIV in Tanzania

**DOI:** 10.1101/2023.11.13.23298478

**Authors:** Makyao Minja, Tusaligwe Mbilinyi, Bryceson Mkinga, Erick G. Philipo, Joyce Owenya, Manase Kilonzi

## Abstract

**Objective:** This study aimed to assess the prevalence of CM, treatment practice, and the associated factor’s post-introduction of Tenofovir Lamivudine and Dolutegravir (TLD) regimen among PLHIV in Tanzania.

**Methods:** This was an analytical cross-sectional study, and the data was collected retrospectively in three public regional referral hospitals (RRHs) in Dar es Salaam, Tanzania. A total of 405 files of the PLHIV admitted in the medical wards on the TLD regimen from January 2019 to December 2022 were reviewed. The collected information includes patient demographic characteristics, Cryptococcal status, CD4 level at the time of CM diagnosis, status of using ART, CM treatment approach, and outcome. Data was analyzed using SPSS software version 23.

**Results:** Out of 405, the majority 267(65.9%) were female, 224(55.3%) were aged between 36 - 55 years, and 293(72.3%) married. ART defaulters were found to be 37(9.1%). The prevalence of CM was found to be 48(11.9%), out of which 42(87.5%) received fluconazole alone. ART defaulter and marital status significantly (p-value < 0.05) were associated with those who tested CM positive.

**Conclusion:** The study found the prevalence of CM among PLHIV to be significantly high and the majority were treated with fluconazole alone. ART defaulters and marital status were significantly associated with one being CM positive. Responsible authorities and stakeholders should enforce guideline adherence and PLHIV should be encouraged on medication adherence.

## I. Introduction

Cryptococcal Meningitis (CM) is one of the leading causes of laboratory-confirmed meningitis in adults living in countries with a high burden of HIV especially in Sub-Saharan Africa (SSA) [1,2]. CM accounts for over 100,000 incident cases of meningitis per year in the SSA [3]. The burden of CM in PLHIV is high and globally is estimated to cause 15%-20% of HIV-related deaths. In low-income countries (LICs) 220,000 cases of CM are estimated per year resulting in 181,000 deaths [4]. In Tanzania, reports have shown that cryptococcosis has 50% fatality at any dose of antifungal given [5]. CM is an opportunistic fungal infection usually affecting immune-compromised hosts like People Living with Human Immune-deficiency Virus (PLHIV), diabetic patients, solid organ transplant recipients, and patients receiving immunosuppressant [6]. The infection is not transferred from person to person, but results from the inhalation of infectious spores mainly found in bird droppings, that multiply in the lungs and can spread to the membranes covering the brain and spinal cord causing meningitis [7]. In HIV-infected patients, the disease is considered as a result of recurrence of latent infection which occurs months to years after the initial exposure event [8,9].

In 2019, Tanzania introduced a Dolutegravir (DTG) based regimen of TLD as the preferred first-line antiretroviral therapy (ART) for most adults and adolescents for HIV-1 infections [10]. From the two landmark clinical trials named ADVANCE and NAMSAL performed in SSA, the efficacy of DTG, and non-emergence of resistance support its use as a first-line regimen for ART-naïve adults with HIV-1 [11]. Patients on the DTG-based regimen had high weight and the viral load was suppressed more quickly compared to the Efavirenz-based regimen [12]. TLD is affordable and well tolerated and improves the clinical outcomes of patients) [13,14]. On the other hand, triple therapy of Amphotericin B, flucytosine (5-FC), and fluconazole is recommended for the treatment of CM. The guidelines recommend two weeks of amphotericin B and 5-FC for the induction phase, followed by fluconazole for the merging and maintenance phases of treatment [15]. Studies report that the use of fluconazole alone is inferior and is associated with high morbidity and mortality compared to triple therapy [16]. However, despite advances in HIV, and CM management, the majority of PLHIV-living LICs face hospital acute mortality in the range between 30-50% [17].

Other factors associated with the burden of CM among PLHIV are reported to be; Being ART naïve and ART defaulter, being male, living in rural areas, being hospitalized, having low CD4 count, and having increasing age with CD4<100 cells, and having body mass index of <18.5kg/m²[18]. Making age, gender, body mass index, and ART adherence major determining factors for cryptococcal infection among PLHIV [19].

Similar to other World Health Organization (WHO) member states, Tanzania adopted the use of a integrase inhibitors-based ART regimen as first-line therapy for adults with PLHIV, and through its guidelines, triple therapy was introduced for the management of CM. Despite the efforts made by the WHO and its member states, the majority of deaths among PLHIV are associated with CM [20]. However, there is a scarcity of information with regard to the prevalence of CM and treatment practice’s post-introduction of integrase inhibitors-based regimens as first-line therapy for PLHIV in LICs. Therefore, this study aims to determine the prevalence, treatment practice, and associated factors of CM among PLHIV post-introduction of TLD in Dar es Salaam, Tanzania.

## II. Materials and Methods

### A. Study design and area

This was a hospital-based cross-sectional study using a retrospective data collection approach. Data were collected from March to June 2023 in public regional referral hospitals (RRHs) in Dares Salaam, Tanzania i.e., Temeke, Amana, and Mwananyamala referral hospitals. The chosen hospitals have Care and Treatment Centers (CTC) that serve the majority of PLHIV residing in Dar es Salaam city and surrounding areas.

### B. Study population

People Living with HIV (PLHIV) admitted to the medical wards on TLD from 2019 to 2022 in the three RRHs (Temeke, Amana, and Mwananyamala).

### C. Sample size and sampling technique

A minimum sample size of 153 PLHIV was estimated using the formula for cross-sectional study (n = p(1-p) z^2^/d^2^). Assuming a population of 7.1% from a previous study on the “burden of serious fungal infection” conducted in Tanzania[6], the confidence level of 95% (z = 1.96) and precision (d) of 5%,. On data collection, a total of 405 PLHIV on TLD admitted to the medical wards from 2019 to 2022 files were retrieved in the three visited RRHs.

### D. Data collection

An Excel spreadsheet was used to collect data from patients’ files, laboratory test results, and Care and Treatment Clinic (CTC) cards where sections involving treatment practice and associated factors were included, as information about associated risk factors, treatment practice, and prevalence was obtained. The information was collected with the assistance of the respected facility’s responsible healthcare worker. On days of data collection data collectors were given files of the HIV patients admitted in medical wards from 2019 to 2022 from the archives. To observe confidentiality names and other information which could identify the participant were not documented.

### E. Data analysis

Data obtained was analyzed by using Statistical Package for Social Science (SPSS) software, version 23. Descriptive statistics was used to summarize socio-demographic and clinical characteristics; the Chi-square test was used to analyze variables associated with the prevalence of CM and treatment outcomes among those who tested CM positive. A p-value below 0.05 was considered statistically significant.

### F. Ethical issues

Ethical clearance was obtained from the Muhimbili University of Health and Allied Sciences (MUHAS)-Research Ethics Committee (REC). The protocol stated clear that the study will retrieve information from patients medical records and the clearance was issued MUHAS-REC: Ref. No. DA. 283/298/01L/602). Permission from the three RRHs administrations (Temeke, Amana, and Mwananyamala) was sought before data collection. Patients’ files were obtained from medical file archives and special identification number was assigned for each file to protect confidentiality of the participants information. Participants name and another information that could identify the participants were not abducted from the files.

## III. Results

### A. Demographic characteristics of study participants

Out of 405 participants, the majority 224 (55.3%) were between 36-55 years followed by 98(24.2%), 18-35 years of age. Regarding employment status, the majority 167(41.2%) were self-employed and 293(72.3%) were married. Besides, most 267(65.9%) of the participants were female and 37(9.1%) were ART defaulter(**table1**).

**Table 1:**
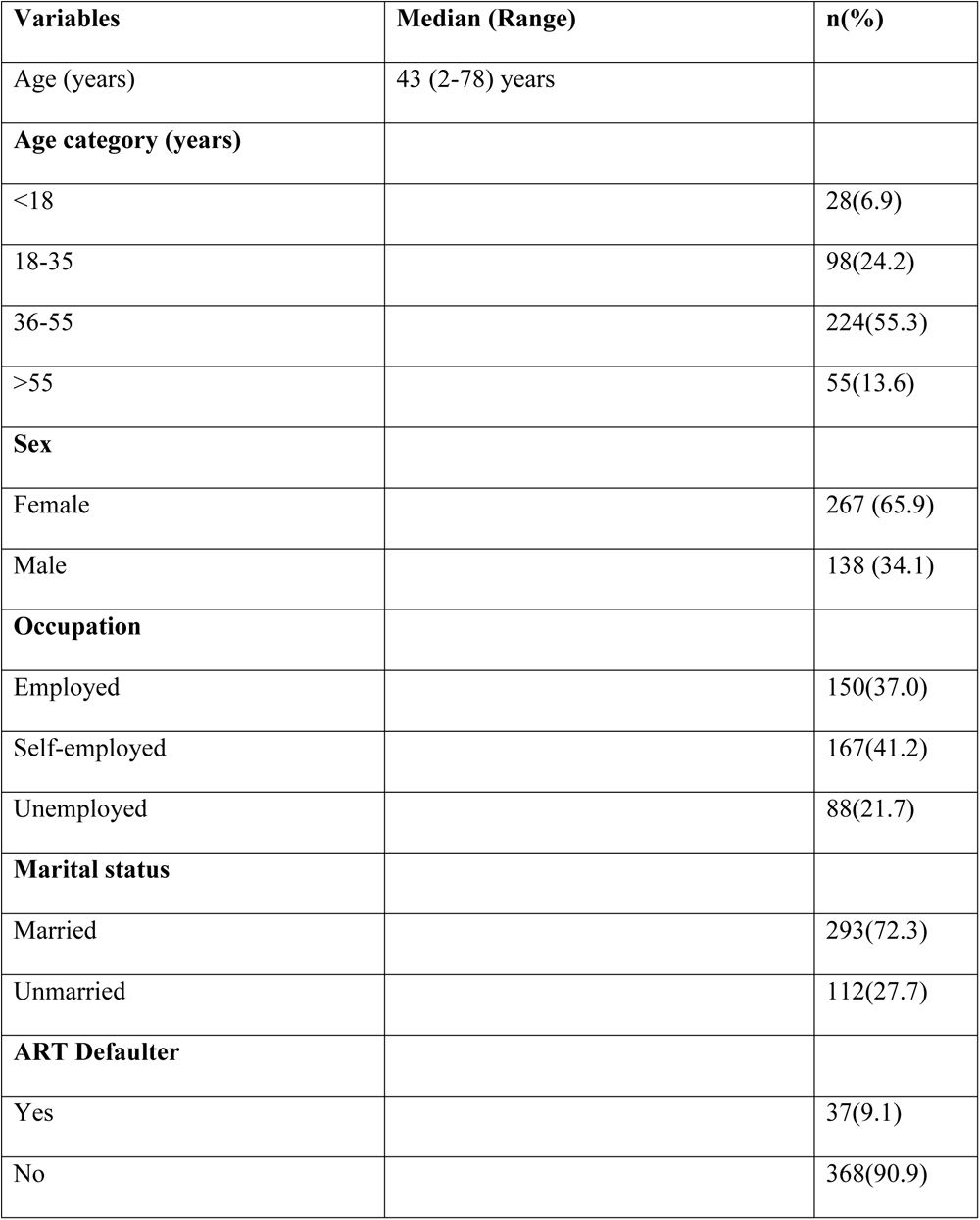
Demographic characteristics of study participants (n=405)

| **Variables** | **Median (Range)** | **n(%)** |
| --- | --- | --- |
| Age (years) | 43 (2-78) years |  |
| **Age category (years)** |  |  |
| <18 |  | 28(6.9) |
| 18-35 |  | 98(24.2) |
| 36-55 |  | 224(55.3) |
| >55 |  | 55(13.6) |
| **Sex** |  |  |
| Female |  | 267 (65.9) |
| Male |  | 138 (34.1) |
| **Occupation** |  |  |
| Employed |  | 150(37.0) |
| Self-employed |  | 167(41.2) |
| Unemployed |  | 88(21.7) |
| **Marital status** |  |  |
| Married |  | 293(72.3) |
| Unmarried |  | 112(27.7) |
| **ART Defaulter** |  |  |
| Yes |  | 37(9.1) |
| No |  | 368(90.9) |

### B. Prevalence of CM, CD4 count at diagnosis, treatment approach and Outcome of treatment among participants

Out of 405 participants, 48 (11.9%) tested CM positive. Of those who tested positive, 6(12.5%) received triple therapy (fluconazole, 5FC, and amphotericin) while 42(87.5%) received fluconazole alone. Following treatment, 4(8.9%) died, 18(40%) recovered, 17(37.8%) were lost to follow-up, 2(4.4%) were given referral, and 4(8.9%) were on going with treatment (**table 2**).

**Table 2:**
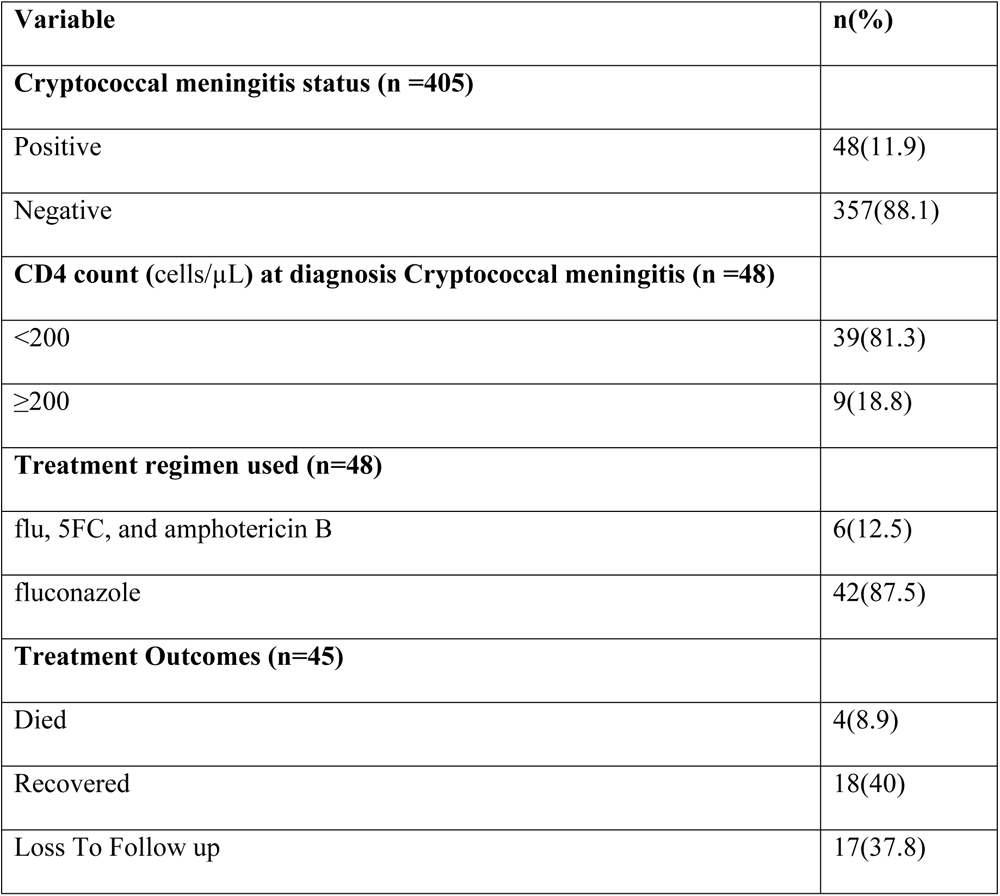
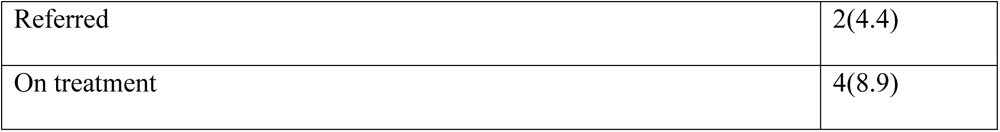
Prevalence of cryptococcal meningitis, CD4 count at diagnosis, treatment approach, and Outcome of treatment among participants (n=405)

| Variable | n(%) |
| --- | --- |
| **Cryptococcal meningitis status (n =405)** |  |
| Positive | 48(11.9) |
| Negative | 357(88.1) |
| **CD4 count (cells/μL) at diagnosis Cryptococcal meningitis (n =48)** |  |
| <200 | 39(81.3) |
| ≥200 | 9(18.8) |
| **Treatment regimen used (n=48)** |  |
| flu, 5FC, and amphotericin B | 6(12.5) |
| fluconazole | 42(87.5) |
| **Treatment Outcomes (n=45)** |  |
| Died | 4(8.9) |
| Recovered | 18(40) |
| Loss To Follow up | 17(37.8) |
| Referred | 2(4.4) |
| On treatment | 4(8.9) |

### C. Factors associated with CM status among study participants

Following Pearson chi-square test, all 37 (100%) ART defaulters tested CM positive compared to non-defaulters; the difference was statistically significant (p-value < 0.01). While those unmarried 24(21.4%) significantly (p-value < 0.01) tested CM positive compared to those who are in marriage 24(8.1%). Other factors including age, sex, and occupation were not associated with being CM-positive among the participants (**table 3**).

**Table 3:** Showing Pearson chi-square test of the demographic characteristics and Cryptococcal meningitis status among participants (n = 405)

| Variables | Cryptococcal meningitis status |  | P-value |
| --- | --- | --- | --- |
| Age (years) | Positive | Negative |  |
| Age category (years) |  |  |  |
| <18 | 0(0.0) | 28(100) | 0.80 |
| 18-35 | 17(17.3) | 81(82.7) |  |
| 36-55 | 25(11.2) | 199(88.8) |  |
| >55 | 6(10.9) | 49(89.1) |  |
| Sex |  |  |  |
| Female | 23(16.7) | 115(83.3) | 0.31 |
| Male | 25(9.4) | 242(90.6) |  |
| Occupation |  |  |  |
| Employed | 8(9.1) | 80(90.9) | 0.64 |
| Self-employed | 20(13.2) | 132(86.8) |  |
| Unemployed | 20(12.1) | 145(87.9) |  |
| Marital status |  |  |  |
| Married | 24(8.2) | 269(91.8) | < 0.01 |
| Unmarried | 24(21.4) | 88(78.6) |  |
| ART defaulter |  |  |  |
| Yes | 37(100) | 0(0.0) | < 0.01 |
| No | 11(3.0) | 357(97.0) |  |

### D. Factors associated with Cryptococcal meningitis treatment outcomes among study participants

Among those who were given a recommended regimen none died, none neither had loss to follow up nor referral, 3(60.0%) recovered and 2(40%) were on going with treatments while among those given fluconazole alone; 4(10%) died, 15(37.5%) recovered, 17(42.5%) were loss to follow up, 2(5%) were referred and 2(5%) are ongoing with treatment and the difference were slightly significant (p-value = 0.048). Other factors like age, sex, occupation, marital status, ART defaulter status, and CD4 count at the time of diagnosis were not statistically related to CM treatment outcome. However, for those who had a CD4 count < 200 cells/µL at the time of diagnosis, 4 (11.1%) died, and 12(33.3%) recovered while those who had a CD4 count of ≥200 cells/µL none died, 6(66.7%) recovered (**table 4**)

**Table 4:**
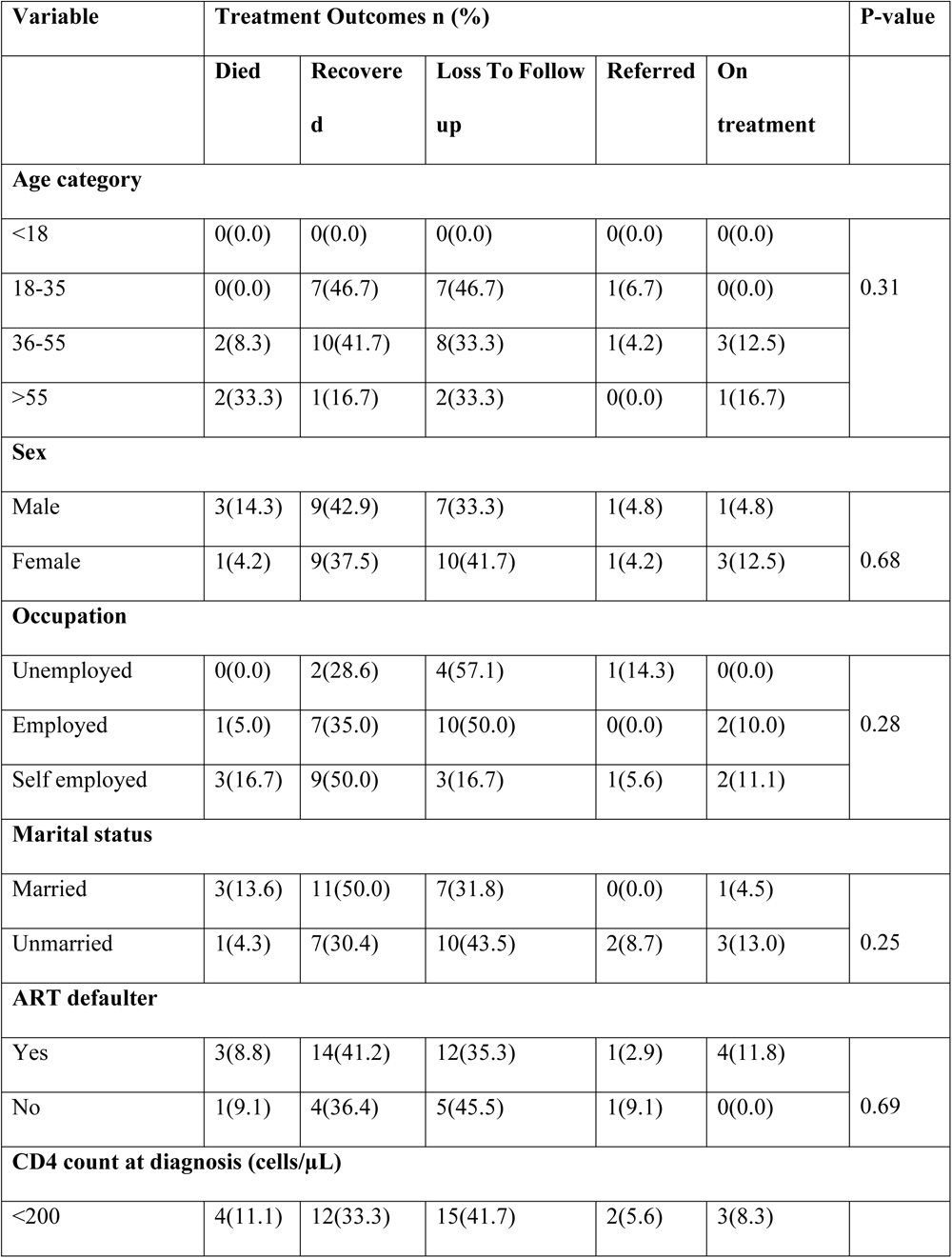
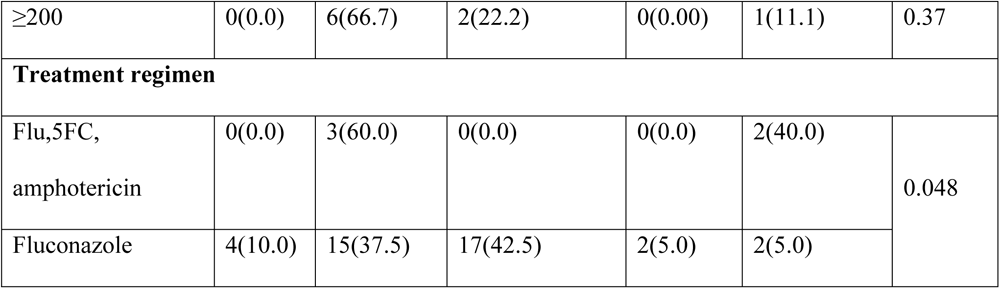
Showing Pearson chi-square test of the demographic characteristics and Cryptococcal meningitis status among participants (n=45)

## IV. Discussion

The objective of this study was to determine the prevalence, treatment, and factors associated with CM post-introduction of TLD regimen among PLHIV in Tanzania. Out of 405 reviewed patient files, the study found that 11.9% had CM, out of which 12.5% received the recommended regimen (triple therapy = Fluconazole, 5-FC, and amphotericin B) and 87.5% received fluconazole alone. Following treatment, 40% recovered while 37.8% were lost to follow-up. ART defaulter and marital status were significantly associated with those who tested CM positive; neither of those treated with triple therapy died, referred, or lost to follow-up compared to those on fluconazole alone.

The prevalence observed in our study is comparable to what was observed in developing countries like Cameroon (11.2%), and Ethiopia (11.4%) [21,22]. The reported prevalence is slightly higher than what was observed in Sierra Leone (7.7%) and Indonesia (7.1%) and higher than what was reported in London (5%), Nigeria (5.1%) and Haiti (1.1%) [23,24]. Varying prevalence’s of CM were observed in different studies could be due to the type of the study design and data collection approaches. Also, some studies included all naive and experienced PLHIV patients while others recruited only those on ART for some time. Our studies assess the CM status of the PLHIV admitted in the hospital for medical reasons which could explain the observed high prevalence. Nevertheless, differences in laboratory protocol, healthcare system, and geographical location of the study settings have been associated with the inconsistency of the reported CM prevalence among PLHIV [25,26]. Since CM is acquired through inhalation of the yeast droplet from the air, weather condition differences may also influence the reported prevalence.

The study demonstrated that only 12.5% of those with CM were treated with the recommended regimen which is a combination of Amphotericin B and 5-FC during the induction course and Fluconazole during the completion phase. The uses of Amphotericin B and flucytosine are reported to reduce CM-related mortality by 40% and clinical trials report that the use of fluconazole alone in the management of CM is inferior [16]. Similar to findings from this study PLHIV with CM treated with fluconazole alone, the majority died or were lost to follow-up compared to those given triple therapy. Despite being recommended by WHO and adopted by member countries like Tanzania, the use of triple therapy in the management of CM in most of the developing countries is low [27]. The poor availability of 5-FC and amphotericin B and the associated cost of purchasing the two medications could be a reason for their poor use in developing countries [28]. The use of a drug like amphotericin B requires laboratory tests prior to and during the time of using the drug in order to prevent patients from toxicity [29]. The latter increases the cost of treatment, taking into consideration that the majority of people in developing countries rely on out-of-pocket mode of payment and the level of poverty could be a reason why healthcare providers (HCPs) opt to use fluconazole alone [30].

In addition, triple therapy involves a different route of drug administration, as fluconazole and 5-FC are given by infusion, and amphotericin B is given intrathecally. The competency of administering amphotericin B among HCPs and the complexity of monitoring for toxicity could be another reason why most HCPs opt for the use of fluconazole alone [31]Training of the HCPs and people involved in the care of PLHIV on how to use triple therapy should be in place to equip HCPs with knowledge and skills on how to dose and administer the medications [28]. Nevertheless, in Tanzania, as in other developing countries patients appear late in the hospital and PLHIV with CM end up admitted and treated as an inpatient through a medical ward. However, medicines and consumables for PLHIV which are purchased and sent to the facilities through a vertical program are available in the CTC. Therefore, proper communication should be initiated to ensure PLHIV admitted in the wards receive medications from CTC. The latter will foster the use of triple therapy as fluconazole, 5-FC, and amphotericin B are grouped as drugs for opportunistic infections (OIs) and are procured through vertical programs [29,32].

The study also observed that ART defaulters were significantly associated with those who tested CM positive. The findings are consistent with what has been reported repeatedly that stopping the use of ART leads to increased viral load, suppressing CD4 cells, and exposing patients to OIs like CM. HIV infection is the common risk of CM, as the risk is estimated to be 95% in PLHIV living in middle-low-income countries and 80% in high-income countries. Despite TLD being effective and user-friendly, current studies still report inadequate adherence to ART among PLHIV [33,34]. Although WHO recommends screening for CM to PLHIV with CD4 levels < 100/ul and naive, studies report the significant prevalence of CM among ART-experienced patients. To fight against the deadly disease, screening for CM among PLHIV should be done regularly despite the CD4 level and years of being on ART [18]. Furthermore, the use of fluconazole as prophylaxis should be scaled up to all PLHIV, particularly in developing countries where the majority have immune problems and access to medical care is a challenge [35]. The latter is possible by taking the example of Isoniazid prophylaxis therapy (IPT) for the prevention of tuberculosis among PLHIV [19].

The study found that the prevalence of CM was higher among unmarried people compared to married people. Several studies report the importance of PLHIV having a close relative/friend or spouse be aware of his/her status [36,37]). Advantages of disclosing HIV status include reducing the level of stigma, having someone to remind a patient to take medication, and clinic schedules and it improves the quality of living [38]. Therefore, more efforts should be implemented to ensure that the majority of PLHIVs disclose the status at least to one of the close spouses/relative or friend. Moreover, our study found no significant difference in CM prevalence between males and females. The findings are not similar to what was reported in other studies in which the prevalence of CM was high in males compared to females [20]. The difference observed could be due to the study setting; our study was conducted in the city of Dar es Salaam, in which both men and women are considered breadwinners of the family [39]. Therefore, being outdoors both men and women are equally exposed to contaminated environments.

## V. Conclusion

The study found the prevalence of CM among PLHIV to be significantly high. Unlike the guideline recommendation, the majority of PLHIV with CM were treated with fluconazole alone instead of triple therapy. ART defaulters and marital status were significantly associated with one being CM positive. Responsible authorities and stakeholders should enforce guideline adherence and a mechanism should be instituted to ensure PLHIV adheres to medication. Taking an example of tuberculosis treatment, in which a victim must visit the clinic with a relative, strategies should be proposed to ensure PLHIV discloses their status to at least a spouse/relative or a close friend.

### VI. List of abbreviation

5-FC: 5-fluorocytosine(Flucytosine)
ART: Antiretroviral Therapy
CTC: Care and Treatment Centre
CD4: Clusters of Differentiation 4
CM: Cryptococcal meningitis
PLHIV: People Living With Human Immunodeficiency Virus
HIV: Human Immunodeficiency Virus
HCPs: Health Care Providers
IPT: Isoniazid Prophylaxis Therapy
LIC: Lower Income Countries
MUHAS: Muhimbili University of Health and Allied Sciences
RRH: Regional Referral Hospitals
SPSS: Statistical Package for Social Science
TLD: Tenofovir Lamivudine Dolutegravir

## Data Availability

All relevant data are within the manuscript and its Supporting Information files.

## VII. Acknowledgements

The authors sincerely thank the Muhimbili University of Health and Allied Sciences – Research and Ethics Committee (MUHAS-REC) for providing ethical clearance and administration of the visited hospitals for providing permission to conduct data collection. We also thank each appointed hospital HCP who helped data collectors to retrieve files and clarified some of the unexplained information.

